# Gender Differences in Health Protective Behaviors During the COVID-19 Pandemic in Taiwan: An Empirical Study

**DOI:** 10.1101/2021.04.14.21255448

**Authors:** Jasmine Tan, Yilin Yoshida, Kevin Sheng-Kai Ma, Franck Mauvais-Jarvis

## Abstract

**Introduction:** Severe acute respiratory syndrome coronavirus 2 (SARS-CoV-2) infection produces more severe symptoms and a higher mortality in men than in women. The role of biological sex in the immune response to SARS-CoV-2 is believed to explain this sex disparity. However, the contribution of gender factors that influence health protective behaviors and therefore health outcomes, remains poorly explored.

**Methods:** We assessed the contributions of gender in attitudes towards the COVID-19 pandemic, using a hypothetical influenza pandemic data from the 2014 Taiwan Social Change Survey. Participants were selected through a stratified, three-stage probability proportional-to-size sampling from across the nation, to fill in questionnaires that asked about their perception of the hypothetical pandemic, and intention to adopt health protective behaviors.

**Results:** A total of 1,990 participants (median age 45.92 years, 49% women) were included. Significant gender disparities (p<0.001) were observed. The risk perception of pandemic (OR=1.28, 95% CI=1.21-1.35, p<0.001), older age (1.06, 95%=1.05-1.07, p<0.001), female gender (OR = 1.18, 95% CI = 1.09□1.27, p<0.001), higher education (OR=1.10, 95% CI=1.06-1.13, p<0.001), and larger family size (OR=1.09, 95% CI=1.06-1.15, p<0.001) were positively associated with health protective behaviors. The risk perception of pandemic (OR=1.25, 95% CI=1.15-1.36), higher education (OR=1.07, 95% CI=1.02-1.13, p<0.05), being married (OR=1.17, 95% CI=1.01-1.36, p<0.05), and larger family size (OR=1.33, 95% CI=1.25-1.42, p<0.001), were positively associated with intention to receive a vaccine. However, female gender was negatively associated with intention to receive a vaccine (OR=0.85, 95% CI=0.75-0.90, p<0.01) and to comply with contact-tracing (OR=0.95, 95% CI=0.90-1.00, p<0.05) compared to men. Living with children was also negatively associated with intention to receive vaccines (OR=0.77, 95% CI=0.66-0.90, p<0.001).

**Conclusion:** This study unveils gender differences in risk perception, health protective behaviors, vaccine hesitancy, and compliance with contact-tracing using a hypothetical viral pandemic. Gender-specific health education raising awareness of health protective behaviors may be beneficial to prevent future pandemics.

## Introduction

Severe acute respiratory syndrome coronavirus 2 (SARS-CoV-2) has caused a global pandemic of coronavirus disease 2019 (COVID-19). Studies across multiple countries have indicated that men present with more severe disease and mortality than women.^1^ As of December, 2020, men accounted for 58 percent of total deaths from COVID-19 globally.^2^ To explain this sex disparity, the role of sex differences in expression of angiotensin-converting enzyme-2 receptor (the entry receptor for SARS-CoV-2), and in immune responses have been proposed.^3^ This sex disparity could also be driven in some parts of the world by social and behavioral determinants, such as higher tendency to tobacco and alcohol use in men compared to women^4^, and differences between men and women in perception and respond to all sorts of risks.^5^

There is a paucity of studies incorporating gender constructs in public health. Sex is characterized by genetics, biological, and physiological traits; while gender, according to the Global Health 50/50 definition, refers to socially constructed norms that impose and determine roles, relationships, and positional power in society.^6^ In particular, the gender role theory proposes that individuals undergo gender socialization, during which role expectations are produced by agents of socialization, such as family, work environment, and cultural environment. For example, women’s greater sensitivity to and lower tolerance to risk may be culturally constructed, and as a consequence a preexisting gender disparity in health-related behaviors could be amplified during a pandemic.

Studies have suggested that sex- and gender are interacting to produce disparities in COVID-19 vulnerability. The initial public health response to COVID-19 involved the promotion of health-protective behaviors, such as home quarantine or mask-wearing.^7^ A previous meta-analysis studying the response to respiratory virus epidemics and pandemics reported that women were 50% more likely than men to practice protecting behaviors, such as mask-wearing^8^. In the early stage of the COVID-19 pandemic, a Japanese study reported that women more frequently practiced social distancing, while men were less likely to adopt preventive strategies^9^. Further, evidence indicates that men exhibit a lower influenza risk perception in the working environment and in clinics than women^10^. Overall, women exhibiting greater health-protective behaviors towards viral infections than men may be attributed to their comparatively higher health-related risk perception, for women more frequently serve as care providers in a family.^11^ In Taiwan, the perception and behavioral responses to contagious diseases, such as COVID-19, including mask wearing, implementing social distancing, contact-tracing and vaccination, have played a vital role in the successful reduction in disease transmission.^12^

The purpose of this study was to evaluate the impact of gender on health-related risk perception and health protective behaviors against a hypothetical influenza pandemic, by analyzing the data from a large-scale nationwide survey. The aims were twofold, i.e., to investigate the characteristics of risk perception and behavioral responses to infectious diseases, and to examine effects of gender and caring responsibility on health protective behaviors.

## Methods

### Participants and Data Collection Procedures

The data used in this study were obtained from the 2014 Taiwan Social Change Survey (TSCS),^19^ a large-scale longitudinal study that tracks the long-term trends of political, economic, social, and cultural changes through national representative survey data collected jointly by the Institute of Sociology and the Centre for Survey Research of Academia Sinica. Respondent was randomly selected by methods of clustering and systematic sampling. Specifically, the 358 townships and cities were separated into seven clusters. The number of target respondents was estimated according to the size of populations in the townships and cities as the primary sampling unit and then in villages and down to individuals. Sampling was weighted by sex, age, urban setting, and education to match the characteristics of the general population of Taiwan.^13^ Responses were recorded in face-to-face interviews by trained interviewers. Follow-up interviews were done by telephone with a random sample of participants to assess validity of the data. One principal investigator trained all interviewers on the health section.

### Assessment of Risk Perception and Health Protective Behaviors

Questionnaire was developed based on existing questionnaire used in studies on risk perception and precautionary behaviors of the general public during outbreaks of SARS and Avian Influenza. ^14,15^ The questionnaire was based on an integrated model to explain health behaviors, including constructs from the Protection Motivation Theory^16^ and the Health Belief Model.^17^ Risk perception is specified as a combination of perceived severity (a person’s belief on how serious contracting the illness would be for him/her) and perceived vulnerability (a person’s perception of the chance that he/she will contract the disease). The Protection Motivation Theory includes another two key constructs, namely response efficacy (a person’s belief in the effectiveness of the preventive measure) and self-efficacy (a person’s level of confidence in his/her ability to perform the preventive measure). Therefore, participants were asked about preventive measures against the new influenza to measure their health protective behaviors.

All items of the questionnaire (Table 1) were rated on a 5□point Likert scale, ranging from 1 (definitely no) to 5 (definitely yes). The total sample size was 2,005, with a response rate of 53%. This study included 1,990 respondents, with no missing data for any of the study variables. Written informed consent was obtained from each respondent. The ethics committees/institutional review boards of the Academia Sinica, Taiwan approved this study and the consent procedure.

**Table 1.**
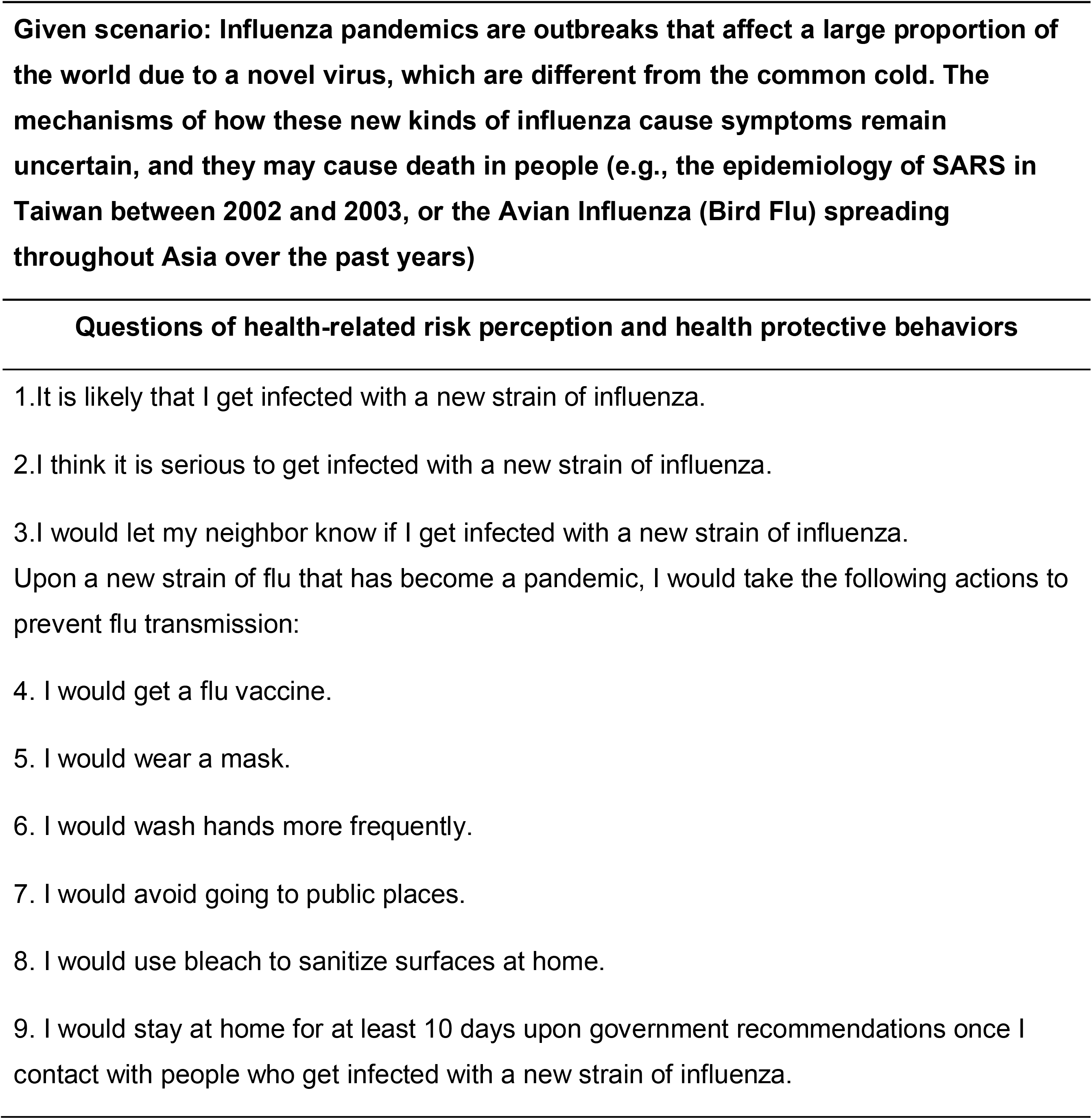
Questions of health-related risk perception and health protective behaviors during a hypothetical pandemic.

### Statistical Analysis

To compare participants’ characteristics by gender, we used chi-square test (Fisher’s exact test when appropriate) for categorical variables and two tailed t test for continuous variables. The preliminary internal structure of questionnaire was explored by using exploratory factor analysis (EFA), a statistical technique to detect common factors of multiple items.^18^Bartlett’s test of sphericity and the Kaiser-Meyer-Olkin measure of sampling adequacy were used to examine whether the correlations of nine items were suitable for exploratory factor analysis. EFA with oblimax rotation was executed, and numbers of factors were decided according to eigenvalues (>1) and Cattell’s scree test.^18^

Variables were then grouped into different dimensions according to the factors found through analysis. Each dimension was treated as a dependent variable for multivariate analysis to examine the difference between men and women. Different adjusted models of socioeconomic demographic factors were derived to examine the interaction effect between these dimensions and health behaviors (i.e., marital status, living condition such as living with parents or/and children, self-related health condition including status of happiness, satisfaction of life, physical health, and education level). Gender stratified analysis was also performed separately. All model-based results are presented with 95% confidence intervals.

## Results

The demographic characteristics of the participants are presented in Table 2. There were 1990 adults aged between 18 and 85 years included in the study. Mean age of the cohort was 45.92 years. The proportion of men and women was equally distributed (50.95% vs 49.05%). More than half the respondents were married or cohabiting (60.90%), with 30.8% aged 20–39 years. Nearly half the respondents were college graduates (45.5%). Socioeconomic characteristics such as marital status, education level, income, self-rated health status and happiness, were significantly different between men and women (p<0.001). More women remained widowed (Mean (SD) =102 (10.5)) than men (Mean (SD) = 16 (1.6)). Men had higher income (27.22% earned more than NT$50000) than women (12.5% earned more than NT$50000). Men also had higher educational attainment than women, with 35.2% versus 29.3% holding university degrees. 2.1% of men and 8.1% of women had no formal schooling. Women reported higher happiness (Mean (SD)=2.87 (0.90)) but poorer physical health status (Mean (SD)=1.99 (0.95)) than men (Mean (SD)=2.68 (1.01) and 2.23 (0.90) respectively).

**Table 2.**
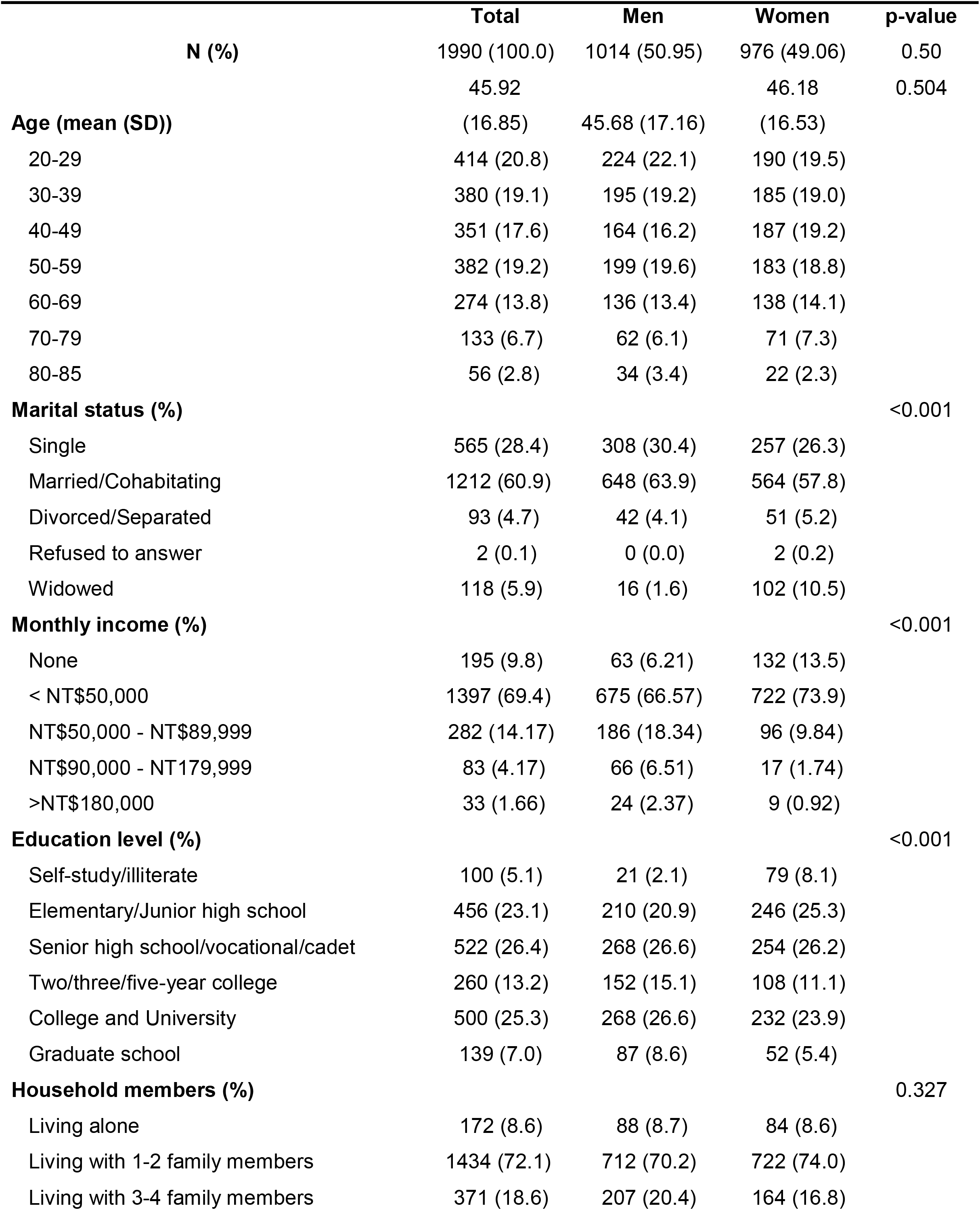
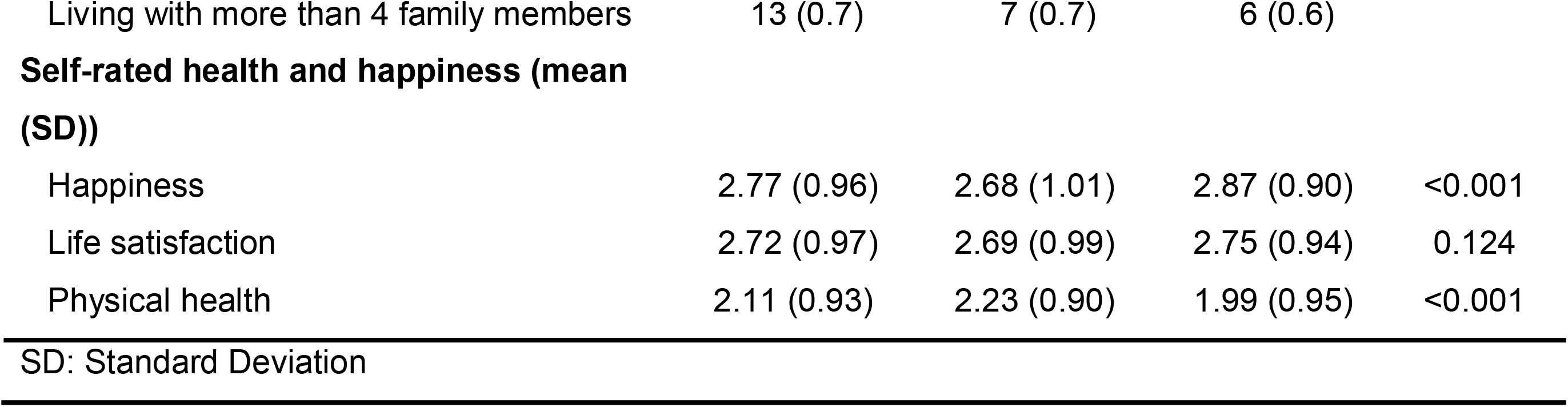
Demographic Characteristics of Participants Stratified by Gender.

Mean score and correlations of each item was presented in Supplementary Table 1. Taiwanese generally showed high intention to adopt preventive measures (mean score >4 in mask wearing, hand wash, and avoidance of public places). The correlations of eight items about risk perception of respondents indicated suitability for EFA (χ^2^ = 2168.56, df = 45, p < 0.001; coefficient of Kaiser-Meyer-Olkin = 0.2521114) (Supplementary Table 1). The EFA of the 9-item questionnaire identified three factors that explained 84.81% of the variance in the data. The rotated factor loadings of structure matrix determined the factor that had the most influence on each variable (Table 3). For example, wearing a face mask (0.78), washing hands (0.75), avoidance of public places (0.73), and sanitization (0.63) showed large positive loadings on factor 1, so we confirmed that this factor described “health protective behavior”. The cut-off value was 0.30.

**Table 3.**
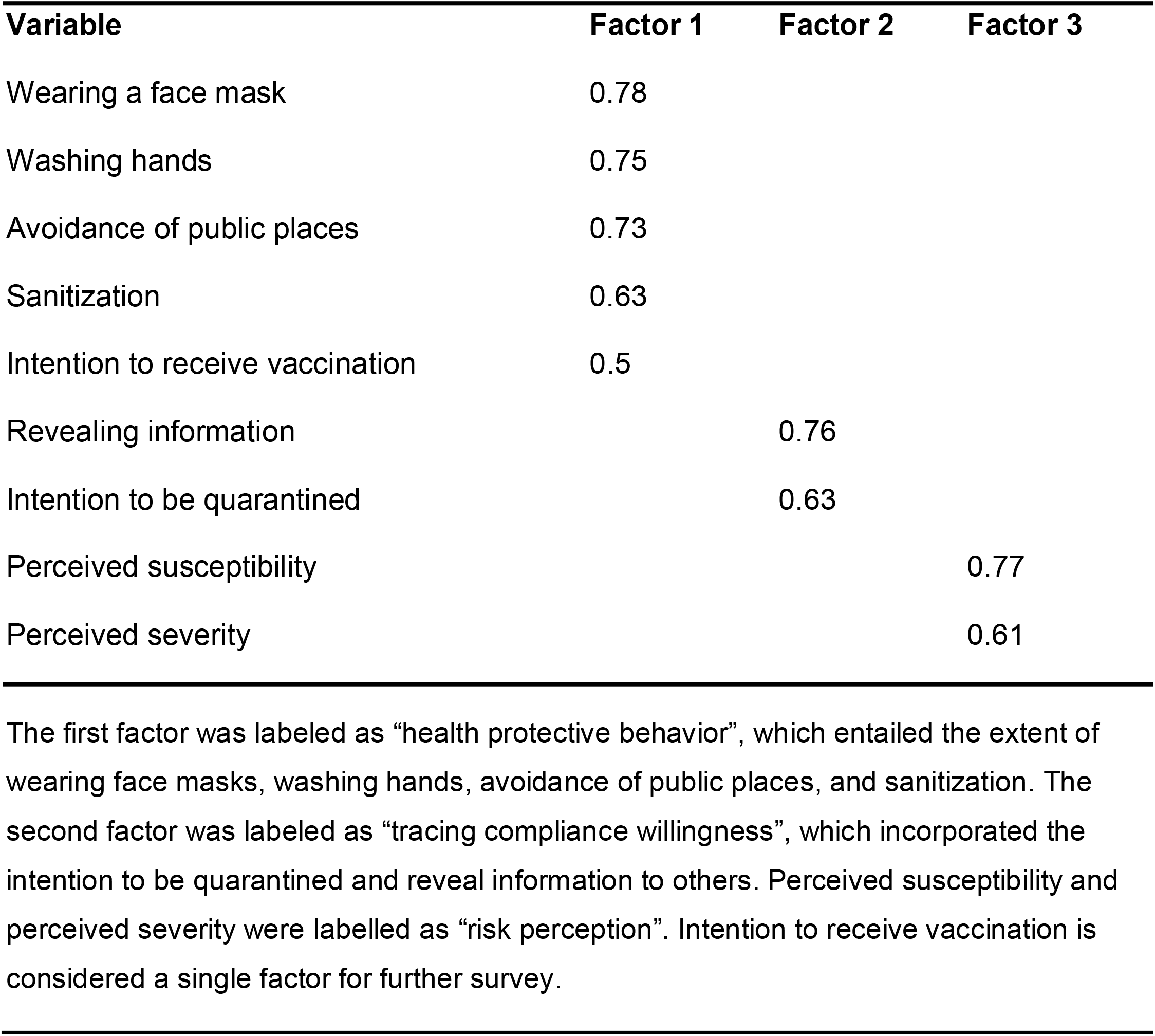
The Rotated Factor Loadings of Structure Matrix.

A four-factor model was chosen as the best analytical dimension based on the factor loadings. The first factor was labeled as “health protective behavior”, which entailed the extent of wearing face masks, washing hands, avoidance of public places, and sanitization. The second factor was labeled as “compliance to contact-tracing”, which incorporated the intention to be quarantined and reveal information to others. The third factor was labelled as “health-related risk perception”, which incorporated perceived susceptibility and perceived severity. Intent to receive vaccination was considered a single factor for further survey.

Figure 1 lists results of the adjusted logistic regression models for factors associated with risk perception, health protective behaviors, intention to receive vaccination, and compliance to contact-tracing, respectively.

**Figure 1.**
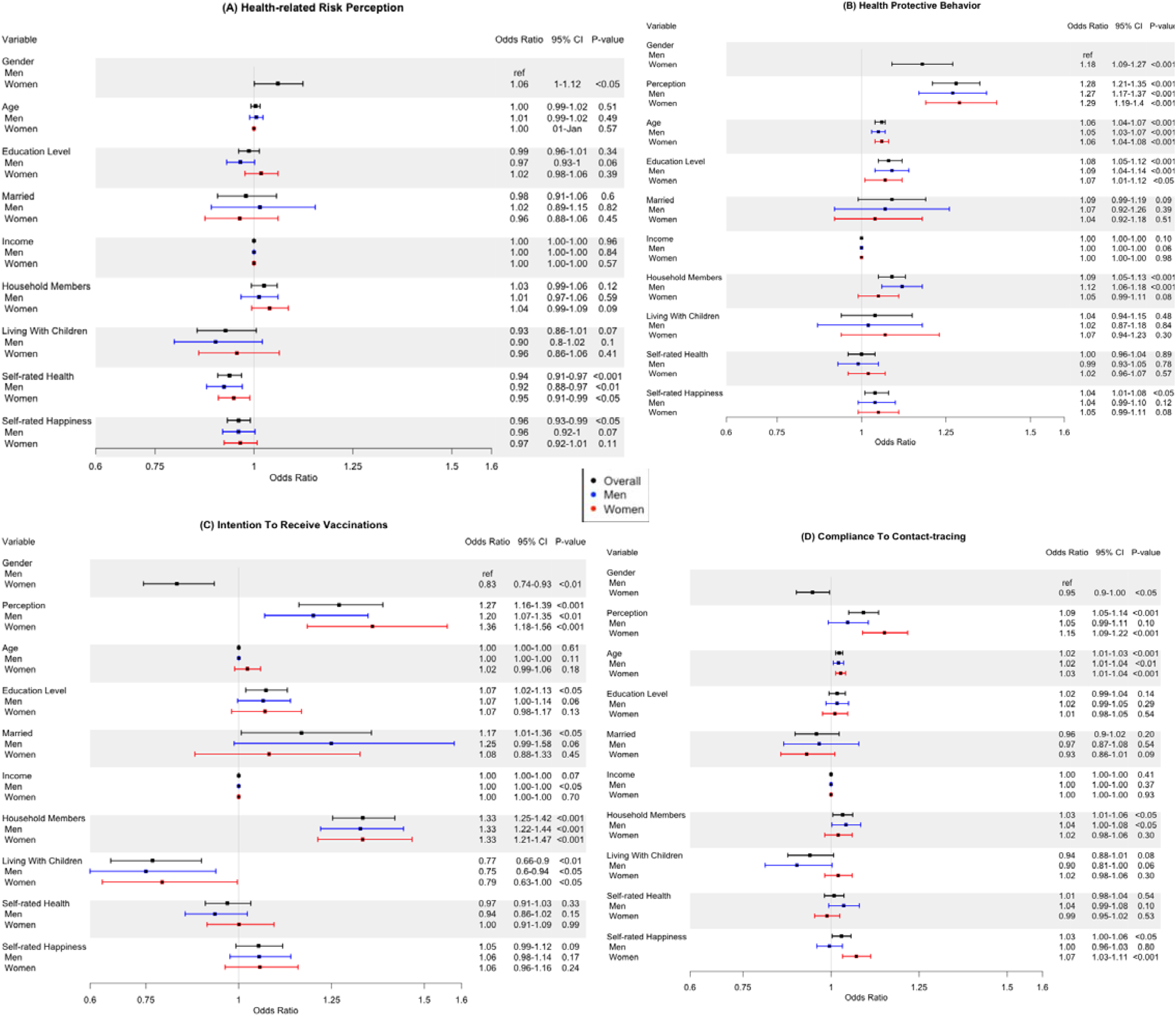
Forest plots showing multivariate logistic regression analysis stratified by gender. Black bar represents result of model among the whole cohort; blue bar and red bar represent men and women respectively. The x-axis represents the odds ratios while the horizontal bars indicated the corresponding 95% confidence intervals. (A) Health-related risk perception (B) Health protective behaviors (C) Intention of receiving vaccination (D) Compliance to contact-tracing.

### Risk perception

Overall, female gender was associated with higher odds of risk perception (OR=1.06, 95% CI=1.00-1.12, p<0.05) compared to male gender. Interestingly, self-rated health (OR=1.06, 95% CI=1.00-1.12, p<0.05) and self-rated happiness (OR=0.96, 95% CI=0.93-0.99, p<0.05) were associated with decreased odds of risk perception (Fig.1A). When stratified by gender, only self-rated health was independently associated with decreased odds of risk perception in both women and men (Fig.1A).

### Health protective behaviors

Women (OR = 1.18, 95% CI = 1.09□1.27, p<0.001), respondents with higher risk perception (OR=1.28, 95% CI=1.21-1.35, p<0.001), older respondents (1.06, 95%=1.04-1.07, p<0.001), respondents with higher education level (OR=1.10, 95% CI=1.06-1.13, p<0.001), and those who lived with household members (OR=1.09, 95% CI=1.05-1.13, p<0.001) were more likely to engage in health protective behaviors (Fig.1B). When data was stratified by gender, age and education level were still independent predictors of health protective behavior in both genders (Fig.1B). However, number of household members was an independent predictor of health protective behavior among men only (OR=1.12, 95% CI=1.06-1.18, p<0.0001) (Fig.1B).

### Intention to receive vaccination

Respondents who exhibited high risk perception (OR=1.27, 95% CI=1.16-1.39, p<0.001), respondents with higher education (OR=1.07, 95% CI=1.02-1.13, p<0.05), married respondents (OR=1.17, 95% CI=1.01-1.36, p<0.05) and respondents with household members (OR=1.33, 95% CI=1.25-1.42, p<0.001) were more likely to exhibit intention to receive vaccination (Fig.1C). Surprisingly, female gender was associated with a decreased intention to receive vaccination (OR=0.83, 95% CI=0.74-0.93, p<0.01) compared to male gender. Similarly, living with children was associated with a decreased intention to receive vaccination (OR=0.77, 95% CI=0.66-0.90, p<0.001) (Fig.1C). Similar results were observed when data was stratified by gender (Fig.1C).

### Compliance with contact-tracing

Female gender was associated with decreased odds of compliance with contact-tracing compared to male gender (OR=0.95, 95% CI=0.90-1.00, p<0.05) (Fig.1D). Older respondents (OR=1.02, 95% CI=1.01-1.03, p<0.001), respondents who lived household members (OR=1.03, 95% CI=1.01-1.07, p<0.05), and respondents exhibiting higher self-rated happiness (OR=1.03, 95% CI=1.00-1.06, p<0.05) expressed higher compliance to contact-tracing compared to their counterparts (Fig.1D). When data was stratified by gender, risk perception (OR=1.15, 95% CI=1.09-1.22, p<0.001) and self-rated happiness (OR=1.07, 95% CI=0.99-1.11, p<0.001) remained as independent predictors of compliance with contact-tracing in women but not in men (Fig.1D). The association between existence of household members and compliance to contact-tracing remained significant in men only (OR=1.04, 95% CI=1.00-1.08, p<0.05).

## Discussion

This study highlights gender disparities in demographic characteristics, health status, as well as adherence to risk perception, and health protective behaviors in a national sample from Taiwan.

The first finding, is that women exhibited a higher perception the of pandemic risk than men. Women were also more likely to adopt health protective behaviors, such as wearing face masks, washing hands, avoiding public places, and practicing sanitization compared to men. Our results are consistent with previous studies reporting that gender influences health behaviors, with women exhibiting higher tendency to adopt new health protective behaviors.^19,20^ Studies from Hong Kong reported that women declared a higher frequency of face mask wearing than men in compulsory situations, such as visiting clinics during flu seasons, and when presenting respiratory symptoms.^21,22^ Published reports during the COVID-19 pandemic also found that age, income, education, and especially gender affect mask-wearing behavior. ^23,24,25^ Women wore masks more often than men,^24^ probably because masks were perceived as a sign of weakness among some men, as suggested by previous work in the United States.^26^ Our findings indicate that women displayed higher level of risk perception and knowledge of preventive measures than men.^25^ Women may be more likely to protect themselves and others by wearing a mask because they handle the majority of caregiving within families^27^, or because of awareness of the preexisting gender inequalities in access to health care that have been further amplified due to the pandemic.^28^

The second finding is that, surprisingly, despite being more likely to adopt health protective behaviors, women, and individual living with children (who were equally distributed across genders) exhibited increased hesitancy to receive vaccination. This observation is consistent with the previous observation that women are more likely to exhibit COVID-19 vaccine hesitancy compared to men.^29,30^ One possible explanation is that women in our study had lower household income than men. Thus, the vaccine acceptability may be lower among these women, if they had perceived the vaccination as an additional expense.^31,32^ Further, women reported lower intention to disclose contact-tracing and to practice home quarantine than men. This may be explained by women’s fear of being discriminated towards infectious disease, which could prevent them from seeking help and medical care.^33^

In subgroup analysis, we observed that living with household members was a predictor of health protective behaviors in men only. However, the positive relationship between living with household members and intention to receive vaccination was observed in both men and women. Respondents bearing family caring responsibilities may explicitly link their fears of the pandemic and their obligations to their household members, and in turn show greater adherence to health protective behaviors.^11^

Our findings may contribute to public health strategies in several ways. First, our research highlights characteristics that may predict compliance to health preventive measures, allowing risk communications to be targeted. In men, interventions should focus on increasing mask-wearing behavior and the importance of adopting nonpharmaceutical preventive measures to protect family members.^34^ In women, individual with children, individuals with lower education level, and individuals who are single or living alone, strategies should focus on decreasing vaccine hesitancy. Finally, women, the elderly, and those living alone, should benefit from education on the importance of contact-tracing and home quarantine that have proven efficient in controlling COVID-19 transmission.^35^

The findings of this study should be considered in the context of certain limitations. First, the study population only included participants from Taiwan, and the findings may not be generalizable to other populations. However, the three stage random sampling procedures, face-to-face interviews and validation of TSCS provided a valuable insight into the whole population residing in Taiwan.^36^ Second, the measurement of household members relied on participants’ characteristics and the questionnaire did not measure the relationship between gender and family, such as by asking men and women separately about whether they felt worried about the effect of pandemic on their household members.

## Conclusion

Our study reveals gender differences in health protective behaviors and vaccine hesitancy. An appreciation of how socio-economic background and gender are influencing health protective behavior could have important implications for public health management and mitigation strategies for future viral pandemics.

## Data Availability

The Center for Survey Research of Academia Sinica in Taiwan is responsible for the data availability and distribution. The dataset is open to the qualified applicants including academic researchers, school faculty, researchers from government organisations, and graduate/undergraduate students.

## Acknowledgement

We thank Da-Wun Sie for comments and suggestions on our study design and statistical analysis.

## Funding

The authors received no specific funding for this work. FMJ was supported by National Institutes of Health Awards DK074970, DK107444, a Department of Veterans Affairs Merit Review Award BX003725, and the Tulane Center of Excellence in Sex-Based Biology & Medicine. YY is supported by a grant (NIH K12HD043451) from the Eunice Kennedy Shriver National Institute of Child Health & Human Development of the Building Interdisciplinary Research Careers in Women’s Health (BIRCWH) Scholar. The content is solely the responsibility of the author and does not necessarily represent the official views of the Eunice Kennedy Shriver National Institute of Child Health & Human Development or the National Institutes of Health.

## Disclosure

These authors declare no conflict of interest.

## Notes

### Competing Interest Statement

The authors have declared no competing interest.

### Author Declarations

The ethics committees/institutional review boards of the Academia Sinica, Taiwan approved this study and the consent procedure.

