## Supplementary Table 1 for "Gender Differences in Health Protective Behaviors During the COVID-19 Pandemic in Taiwan: An Empirical Study"

| **Supplementary Table 1. Mean, Standard Deviation, and Correlation Matrix of ten variables measured** | | | | | | | | | | | | |
| --- | --- | --- | --- | --- | --- | --- | --- | --- | --- | --- | --- | --- |
|  | **Variables** | **Mean** | **SD** | **1** | **2** | **3** | **4** | **5** | **6** | **7** | **8** | **9** |
| 1 | Perceived susceptibility | 2.3103 | 0.9118 | 1 | 0.06* | -0.01 | 0.08** | 0.09*** | 0.07** | 0.03 | 0.04 | -0.01 |
| 2 | Perceived severity | 4.2223 | 0.8353 | 0.06* | 1 | 0.08** | 0.1*** | 0.17*** | 0.12*** | 0.11*** | 0.12*** | 0.09*** |
| 3 | Revealing information | 2.7372 | 0.7066 | -0.01 | 0.08** | 1 | 0.07** | 0.06* | 0.09*** | 0.1*** | 0.1*** | 0.2*** |
| 4 | Vaccination | 3.9132 | 1.2412 | 0.08** | 0.1*** | 0.07** | 1 | 0.31*** | 0.27*** | 0.25*** | 0.21*** | 0.11*** |
| 5 | Wearing a face mask | 4.0253 | 1.2956 | 0.09*** | 0.17*** | 0.06* | 0.31*** | 1 | 0.6*** | 0.46*** | 0.35*** | 0.22*** |
| 6 | Washing hands | 4.5472 | 0.7117 | 0.07** | 0.12*** | 0.09*** | 0.27*** | 0.6*** | 1 | 0.44*** | 0.32*** | 0.22*** |
| 7 | Avoidance of public places | 4.2394 | 0.9689 | 0.03 | 0.11*** | 0.1*** | 0.25*** | 0.46*** | 0.44*** | 1 | 0.41*** | 0.27*** |
| 8 | Sanitization | 3.708 | 1.2313 | 0.04 | 0.12*** | 0.1*** | 0.21*** | 0.35*** | 0.32*** | 0.41*** | 1 | 0.19** |
| 9 | Intention to be quarantined | 3.4326 | 0.7216 | -0.01 | 0.09*** | 0.2 | 0.11*** | 0.22*** | 0.22*** | 0.27*** | 0.19*** | 1 |
| *p <0.05; **p <0.01; ***p<0.001 | | | | | | | | | | | | |

**Data collection procedure**

Our sampling frame consists of registered non-institutional residents aged 18 or over in Taiwan. The sampling area includes basically the main island of Taiwan, excluding the outlying islands. The stratified three-stage probability proportional to size sampling was used to select respondents. An experts’ committee was formed to evaluate reliability of the questionnaire by taking previous survey models and published studies as reference. Preliminary data was collected in April, 2014 to access participants’ perceptions about the survey itself (e.g., length, clarity of questions) and modify accordingly. Data were collected from July to October in 2014 by face-to-face interview. We sent postal letters to all sampled individuals for advanced contacts. Interviewers must follow certain rules when they approached an address. Visits were made at different times of day and on different days of the week. Interviewers were required to make at least three visits before they stopped approaching an address. About two percent of interviews were supervised, and 53% of full productive interviews were back-checked (supervisors check later to see whether interviews were conducted). The total number of issued sample is 4488 (gloss sample size). With 1,927 completed interviews (net sample size), the survey generated a response rate (net sample size/gloss sample size) of 42.94%.

**Sampling Procedures**

The following variables are used to stratify the population frame into seven levels of regions: population density, educational level, the proportion of population over age 65, the proportion of population between ages 15 and 64, the proportion of industrial employment as the total employment, and the proportion of service sector employment as the total employment. The sampling design has three stages. For the first stage, the number of target respondents is decided for each of the seven strata of regions proportionate to the size of their populations. For the second stage, the number of townships is decided for each regional level and is randomly selected from each level. Districts or villages then are randomly selected from each chosen townships. For the third stage, around 21 to 30 individuals ages 18 or over are randomly selected from household registers in each precinct or village.

**Gender effect on marginal model of health-related risk perception and health-related behaviors**

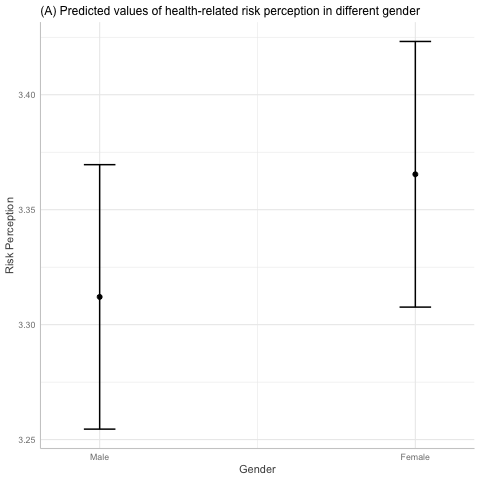

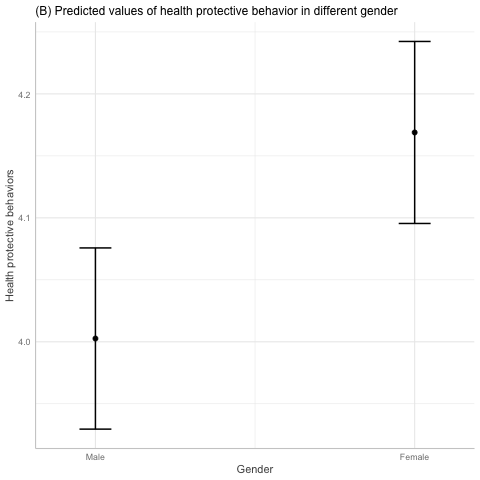

The average effects of both models were derived from the multivariate logistics regression in Figure 1. Women exhibited more health-related risk perception and health-related behaviors than men, assessed by the predicted average marginal effect of health-related risk perception and health-related behaviors. The average effect of health-related risk perception was 3.25 (95%CI, 3.21-3.29) for men and 3.30 (95%CI, 3.26-3.34) for women. Women was also an independent predictor of health-related behaviors, with predicted values of 4.02(95%CI, 3.97-4.07) and 4.19 (95%CI, 4.14-4.24) for men and women, respectively.
